# Clinical Characteristics of 20,662 Patients with COVID-19 in mainland China: A Systemic Review and Meta-analysis

**DOI:** 10.1101/2020.04.18.20070565

**Authors:** Chong Tang, Keshi Zhang, Wenlong Wang, Zheng Pei, Zheng Liu, Ping Yuan, Zhenpeng Guan, Jin Gu

## Abstract

Coronavirus disease 2019 (COVID-19) is a global pandemic and has been widely reported; however, a comprehensive systemic review and meta-analysis has not been conducted. We systematically investigated the clinical characteristics of COVID-19 in mainland China to guide diagnosis and treatment. We searched the PubMed, Embase, Scopus, Web of Science, Cochrane Library, bioRxiv, medRxiv, and SSRN databases for studies related to COVID-19 published or preprinted in English or Chinese from January 1 to March 15, 2020. Clinical studies on COVID-19 performed in mainland China were included. We collected primary outcomes including signs and symptoms, chest CT imaging, laboratory tests, and treatments. Study selection, data extraction, and risk of bias assessment were performed by two independent reviewers. Qualitative and quantitative synthesis was conducted, and random-effects models were applied to pooled estimates. This study is registered with PROSPERO (number CRD42020171606). Of the 3624 records identified, 147 studies (20,662 patients) were analyzed. The mean age of patients with COVID-19 was 49.40 years, 53.45% were male, and 38.52% had at least one comorbidity. Fever and cough were the most common symptoms, followed by fatigue, expectoration, and shortness of breath. Most patients with COVID-19 had abnormal chest CT findings with ground glass opacity (70.70%) or consolidation (29.91%). Laboratory findings shown lymphopenia, increased lactate dehydrogenase, increased infection-related indicators, and fibrinolytic hyperactivity. Antiviral therapy, antibiotic therapy, and corticosteroids were administered to 89.75%, 79.13%, and 35.64% of patients, respectively. Most clinical characteristics of COVID-19 are non-specific. Patients with suspected should be evaluated by virological assays and clinically treated.

## Introduction

Severe acute respiratory syndrome coronavirus 2 (SARS-CoV-2) is the seventh human coronavirus identified, with bats thought to be the original host.^1-3^ SARS-CoV-2 is approximately 50% genetically identical to Middle East respiratory syndrome coronavirus (MERS-CoV) and approximately 79% identical to severe acute respiratory syndrome coronavirus (SARS-CoV) and shows a similar receptor-binding domain structure.^4^ Patients infected with SARS-CoV-2 may be asymptomatic or have mild to severe pneumonia. The syndrome of clinical symptoms caused by SARS-CoV-2 is named as coronavirus disease 2019 (COVID-19).^5^

COVID-19 is the third type of zoonotic coronavirus disease after SARS and MERS occurring in the last two decades. COVID-19 is highly contagious through respiratory droplets and contact and has caused global pandemic. As of April 17, 2020, COVID-19 has spread to 213 countries and regions globally, causing 2,074,529 confirmed cases and 139,378 deaths.^6^ At present, the United States and Europe have become the hardest-hit regions by COVID-19, with 632,781 confirmed cases in the United States of America, 1,050,871 confirmed cases in European region.

Governments worldwide must urgently need to learn from China’s experience, as China experienced a rapid growth period followed by a stable decline period of the COVID-19 epidemic, and currently a recovery period.

Since the outbreak of COVID-19, a large number of articles have been published or preprinted reporting its epidemiologic and clinical characteristics. However, few large studies have been reported, and variations in reporting descriptive data may lead to the misunderstanding of the clinical features of COVID-19. In this review, we systematically investigated the epidemiologic, characteristics, chest computed tomography (CT) imaging, laboratory findings, and treatments of COVID-19 in mainland China. Our findings provide important guidance on the diagnosis and treatment of the current global COVID-19 pandemic.

## Methods

### Search strategy and selection criteria

For this systematic review and a meta-analysis, we searched the PubMed, Embase, Scopus, Web of Science, Cochrane Library, bioRxiv, medRxiv, and SSRN electronic databases for papers published or preprinted in English or Chinese from January 1 to March 15, 2020 with the following search terms: “coronavirus disease 2019” OR “COVID-19” OR “SARS-CoV-2” OR “2019-nCoV” OR “Novel Coronavirus” OR “Wuhan pneumonia”. Additional studies were identified by manually searching the reference lists of primary studies and review articles. The authors of full papers were contacted for additional information when required.

The inclusion criteria were as follows: clinical studies of COVID-19; studies performed in mainland China; number of cases ≥10; and primary outcomes including signs and symptoms, chest CT imaging, laboratory tests, and treatments were available. We excluded studies conducted outside of China; case reports, case series, reviews, abstract, and opinions; those without primary outcomes.

Two investigators (CT, KZ) independently reviewed the title and abstract of the retrieved articles to select eligible articles according to the inclusion criteria. After excluding duplicated and irrelevant studies, the full-text of the remaining studies was reviewed to assess the eligibility for inclusion. The inter-rater agreement of study selection was measured using the κ statistic. If multiple studies used the same dataset or cohort, we included the most comprehensive study with the largest number of participants and excluded the others. Disagreements were resolved by consensus or were arbitrated by a third investigator (ZL).

### Data analysis

Two investigators (CT, WW) independently extracted primary and secondary outcomes from the eligible studies using a predefined and standardized data extraction checklist, with disagreements resolved by discussion. The following information were recorded: first author, province, city, sample size, gender, age, smoker, comorbidities, symptoms, chest CT imaging, laboratory findings, complications, treatments, and prognosis. For missing data, we evaluated online supplementary appendixes or contacted the first or corresponding author to obtain the missing data or more information. If no response was obtained, we calculated the data using methods such as imputing the mean and standard deviation (SD) from the median, interquartile range (IQR), and full range as described by Wan et al.^7^ and Luo et al.^8^

Two investigators (ZP, PY) independently assessed the risk of bias in the eligible studies, with disagreements resolved through discussion. The Risk Of Bias In Non-randomised Studies - of Interventions (ROBINS-I) was used to assess study quality.^9^ ROBINS-I contains seven domains: (1) bias due to confounding factors; (2) bias in the selection of participants for the study; (3) bias in classification of interventions; (4) bias due to deviations from the intended interventions; (5) bias due to missing data; (6) bias in the measurement of outcomes; and (7) bias in the selection of reported results. The categories for risk of bias judgement were “Low risk”, “Moderate risk”, “Serious risk”, “Critical risk”, and “No information”, with a high-quality study defined as one showing a “Low risk” or “Moderate risk”. Studies were excluded when the quality was “Critical risk” or “No information”.

R version 3.6.2 and Stata version 15 (StatCorp, College Station, TX, USA) were used for statistical analyses. All data are expressed as overall summary estimates and 95% confidence intervals (CIs). Cochran’s Q test and I^2^ statistic were used to assess heterogeneity, with p < 0.05 for Cochran’s Q test or I^2^ >50% indicating significant heterogeneity, in which cases the random-effects model was used. Otherwise, the fixed-effects model was used. Heterogeneity was assessed by meta-regression of province, sample size, and age. Potential publication bias was assessed by quantitative Egger’s linear regression test. This study is registered with PROSPERO (number CRD42020171606).

## Results

A total of 3624 records were retrieved, after removing the duplicates, 2751 records were retained. We screened the titles and abstracts and excluded 2493 ineligible records. The full-texts of the remaining 258 records were assessed for eligibility, of which 111 were excluded. Of the selected 147 articles, 143 were written in English and 4 were in Chinese. The study selection flow is shown in Figure 1 and the characteristics on eligible articles is shown in Table 1.

**Table 1.**
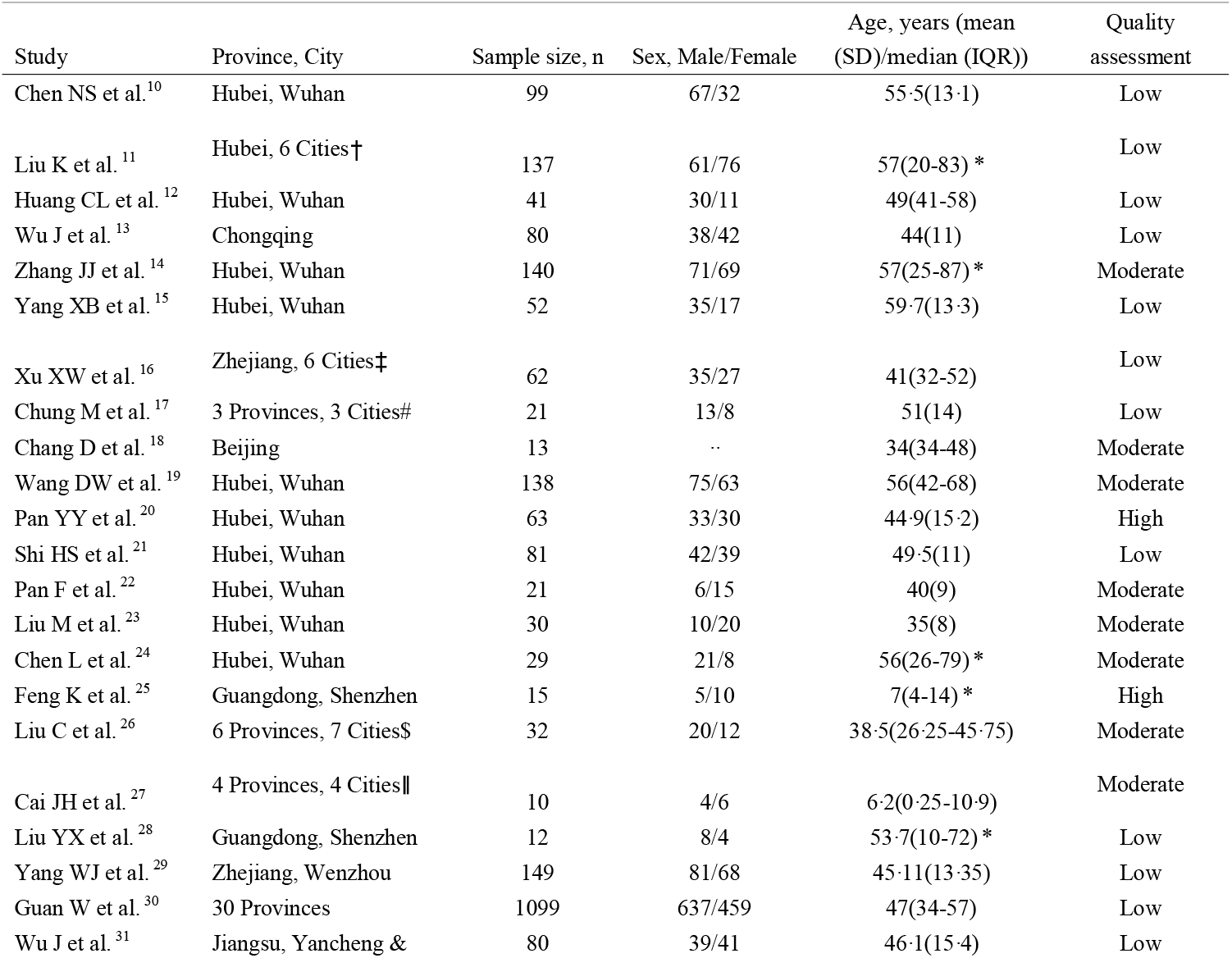
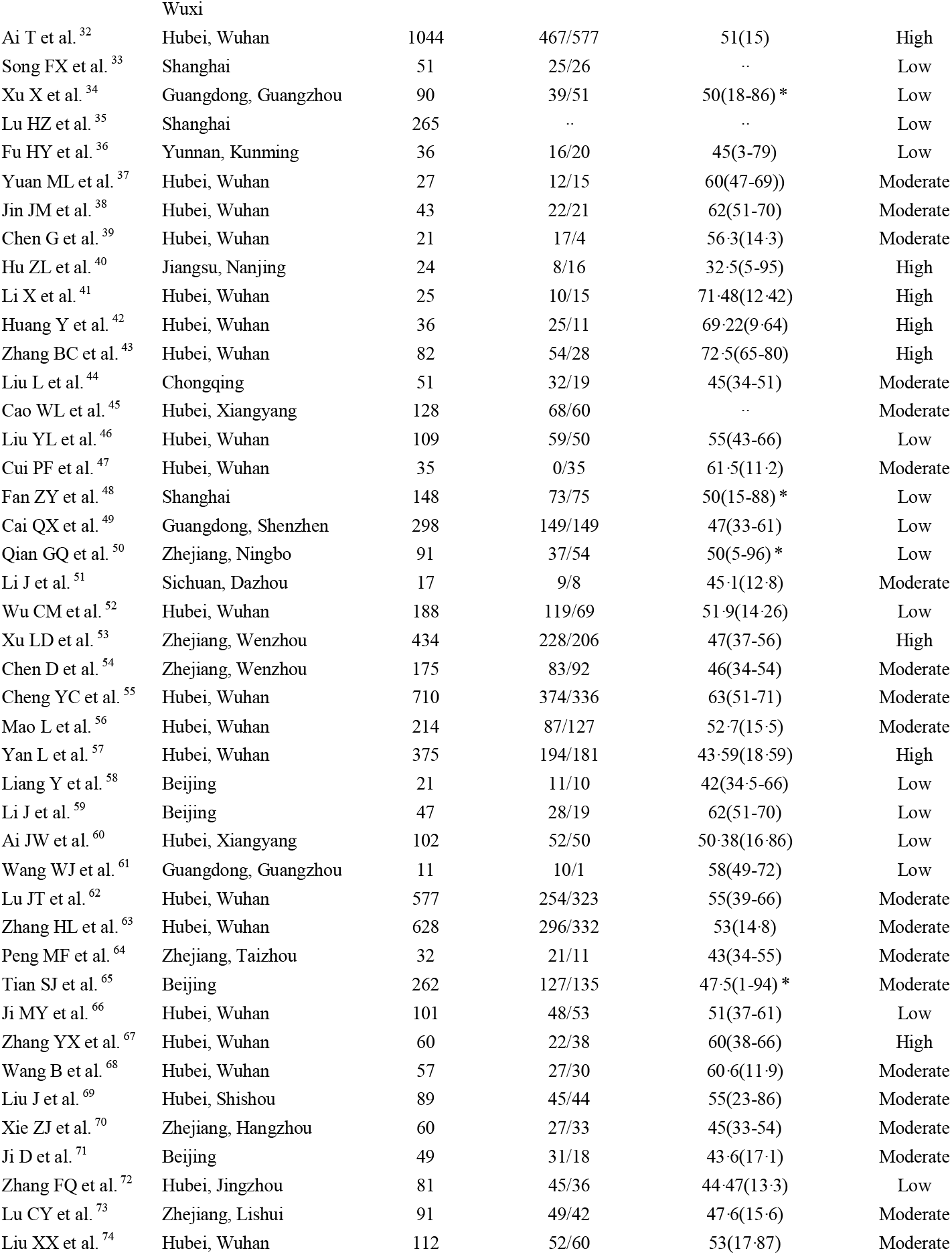
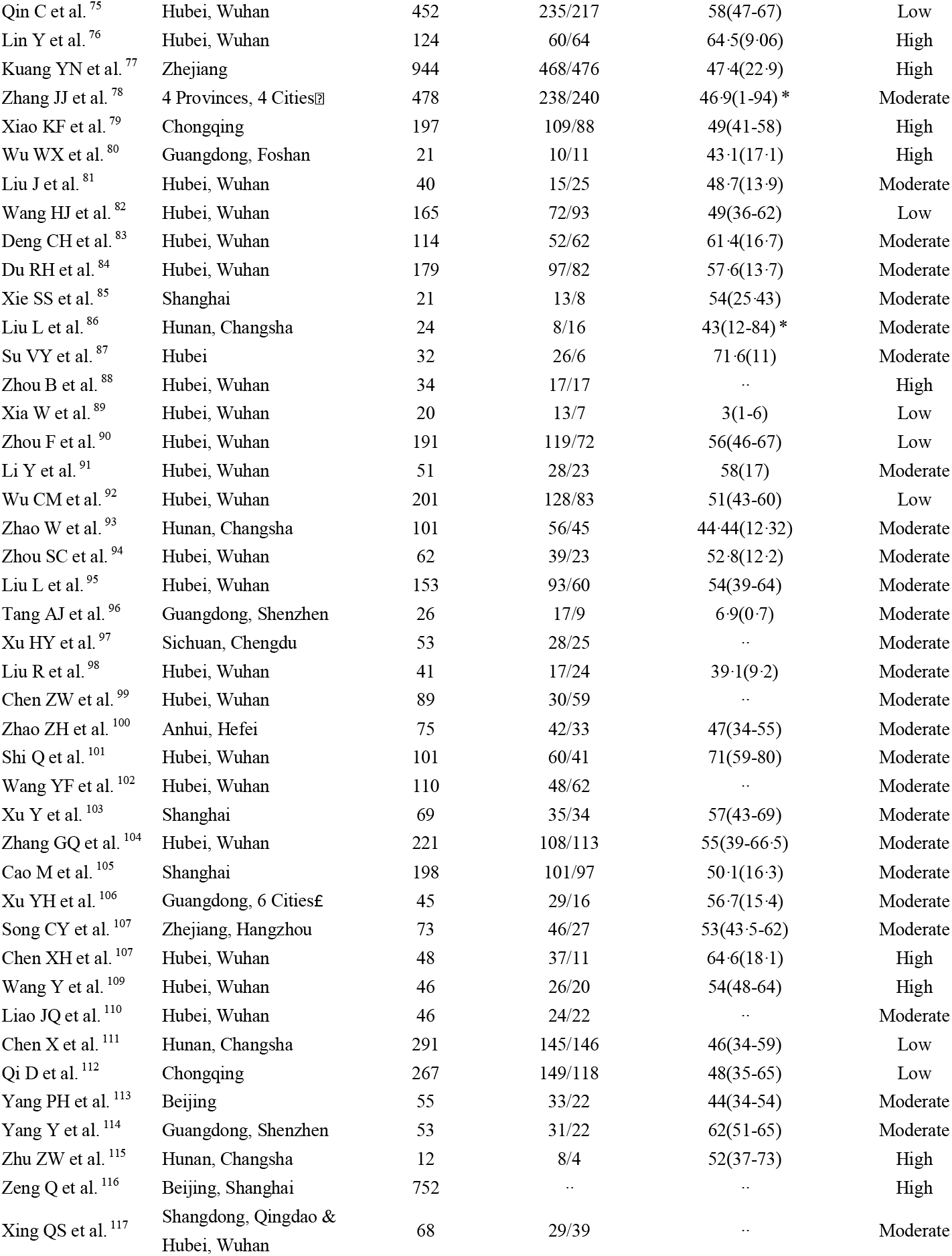
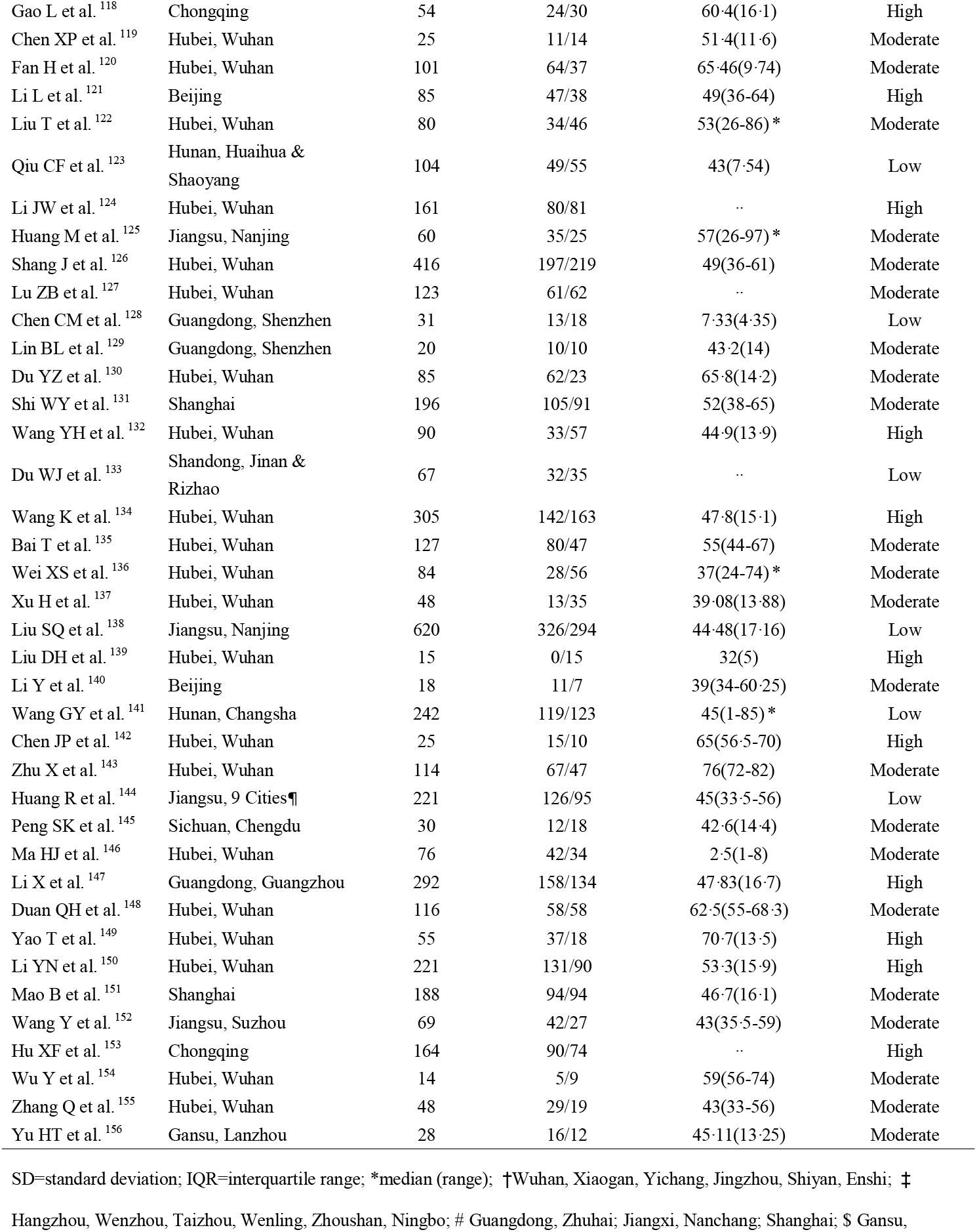
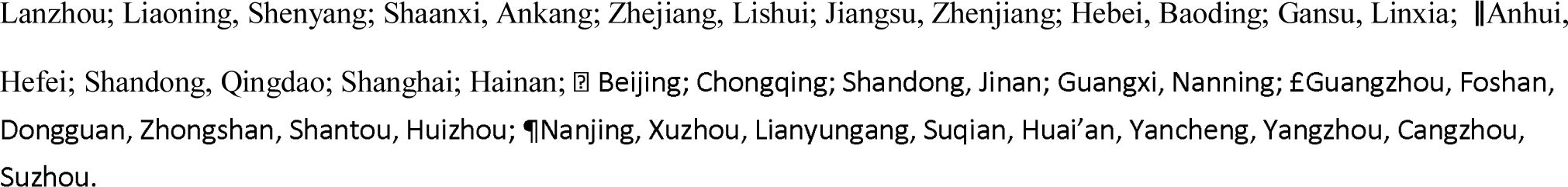
Characteristics of included studies

**Figure 1.**
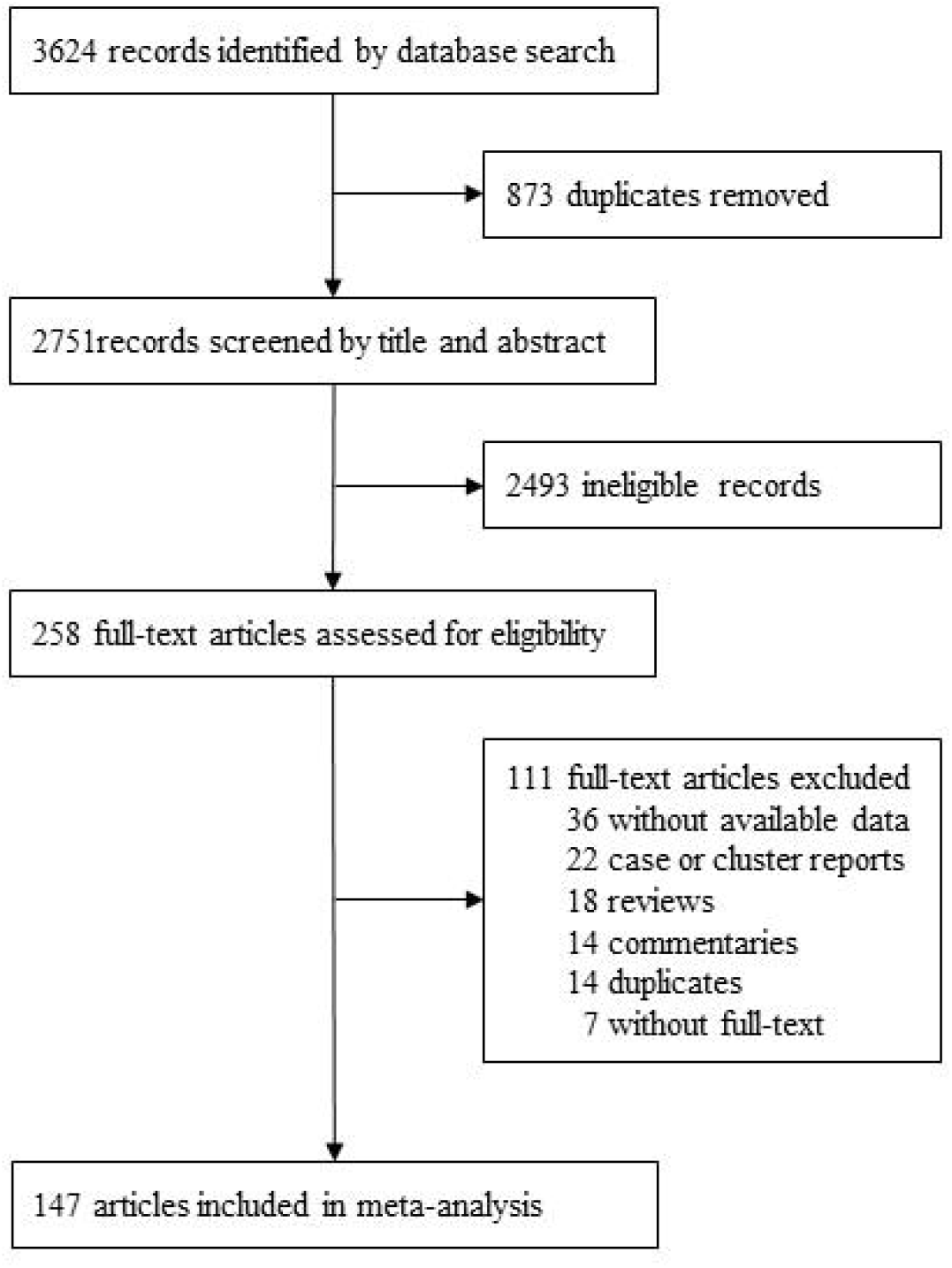
Flowchart of study selection.

The results of quality assessment of the included studies are shown in the appendix (pp 2-7). Overall, 117 high-quality studies and 30 fair-quality studies were included in the meta-analysis.

### Epidemiology and comorbidities of COVID-19

A total of 20,662 patients with COVID-19 were included. Patient ages ranged from 1 month to 100 years old, with a mean age of 49.40 years (95%CI of 45.15–53.65 years). The proportion of males was 53.45% (95%CI 52.02–54.92%). Among 5094 patients in 35 studies, 511 were smokers, accounting for 9.93% of patients (95%CI 7.87–12.54%).

Of 8028 patients with COVID-19, 3066 (38.52%, 95%CI 34.17–43.42%) had at least one comorbidity, including hypertension (20.78%), diabetes (12.04%), cardiovascular disease (8.58%), cerebrovascular diseases (6.01%), chronic obstructive pulmonary disease (4.20%), chronic liver disease (4.19%), malignancy (3.92%), and chronic renal disease (3.11%). (see Figure 2, Appendix pp 8)

**Figure 2.**
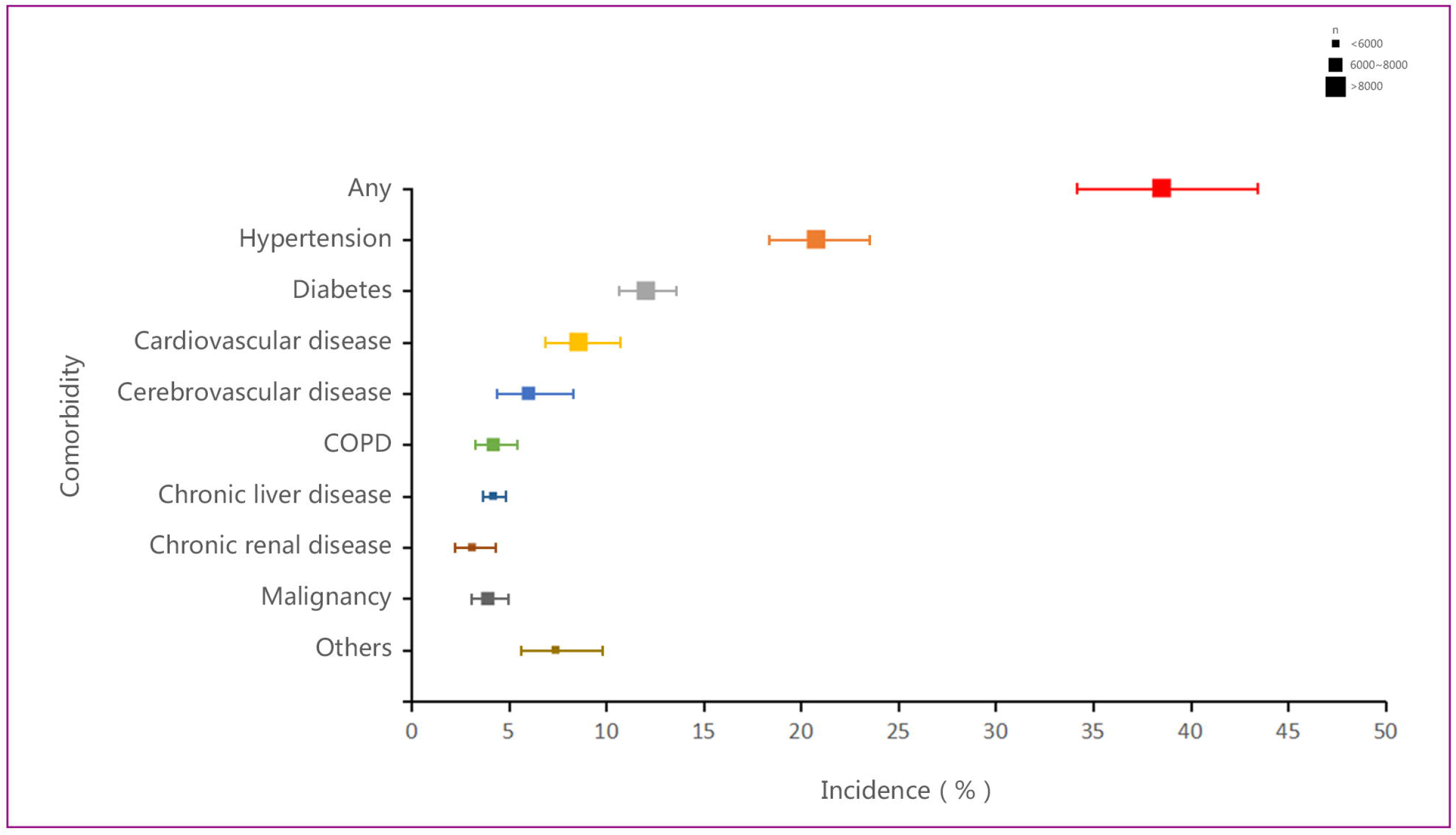
Estimates of the incidence of comorbidities for COVID-19 patients. Note: COPD= chronic obstructive pulmonary disease.

### Signs and symptoms of COVID-19

The most common symptom of COVID-19 was fever. Of 18,513 patients with COVID-19, 13,707 (78.49%, 95%CI 76.47–80.56%) had fever. The proportions of patients with mild to moderate fever were 30.96% and 35.02%, and only 11.45% of patients had high fever. Patients also experienced cough (55.49%, 95%CI 52.71–58.41%), fatigue (30.42%, 95%CI 27.20–34.03%), expectoration (26.64%, 95%CI 24.13–29.41%), shortness of breath (21.07%, 95%CI 16.33–27.17%), dyspnea (17.08%, 85%CI 14.15–20.60%), myalgia (17.06%, 95%CI 14.32–20.32%), chest tightness (14.19%, 95%CI 10.61–18.98%), anorexia (13.78%, 95%CI 10.42–18.21%), chill (12.86%, 95%CI 10.47–15.79%), and headache (10.14%, 95%CI 8.84–11.63%). Less common symptoms were pharyngalgia (9.17%), dizziness (8.92%), diarrhea (8.61%), rhinorrhea (5.21%), nausea (4.56%), vomiting (4.02%), chest pain (3.98%), abdominal pain (3.71%), and hemoptysis (2.62%). (see Figure 3, Appendix pp8-9)

**Figure 3.**
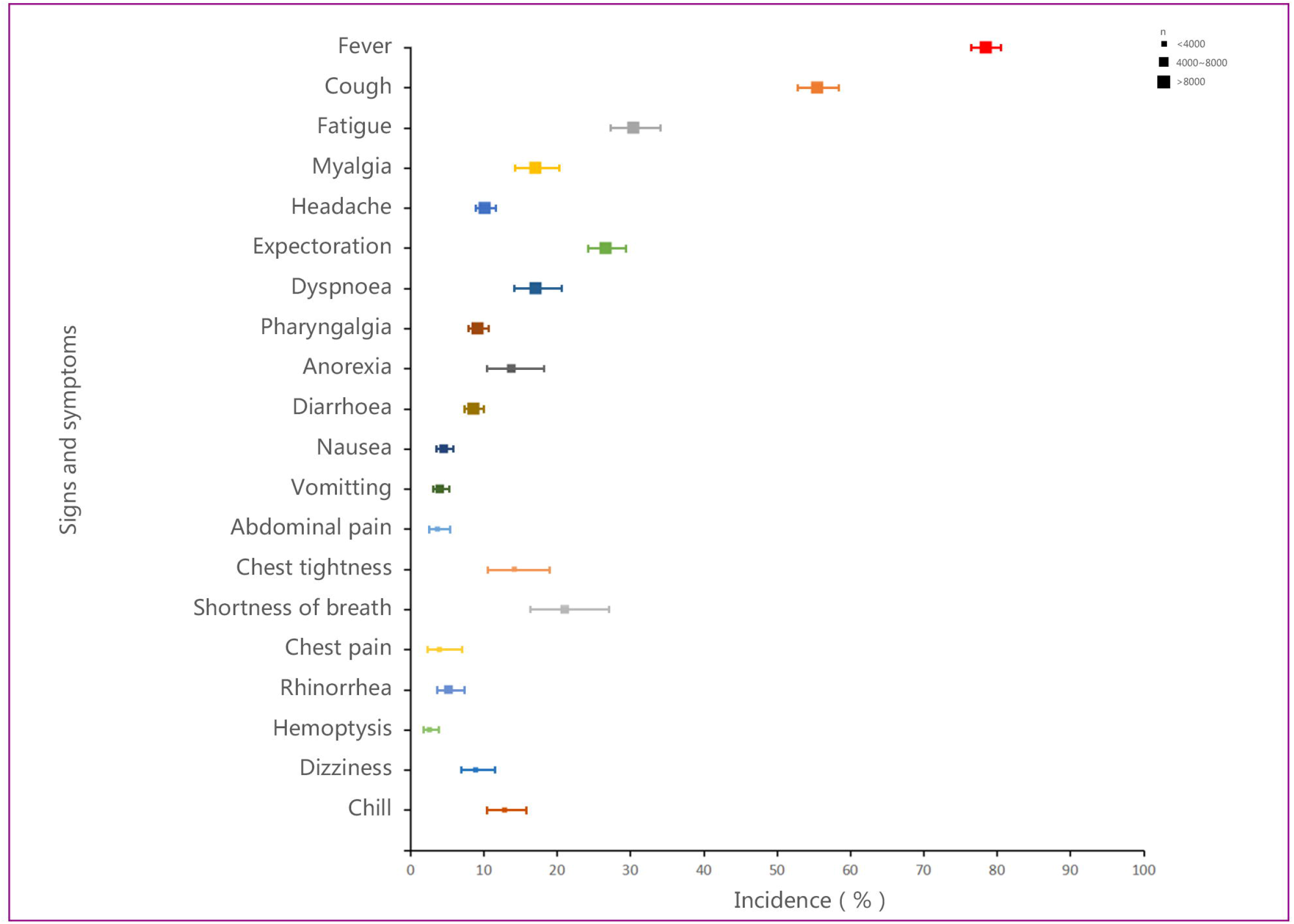
Estimates of the incidence of signs and symptoms for COVID-19 patients.

### CT imaging and laboratory findings of COVID-19

Of 8711 patients with COVID-19, 7919 (95.31%) had abnormal presentations on chest CT imaging, with 74.72% bilateral infiltration, 73.60% peripheral distribution, 70.70% ground glass opacity, 29.91% consolidation, and 8.06% pleural effusion. (Figure 4, Appendix pp 9)

**Figure 4.**
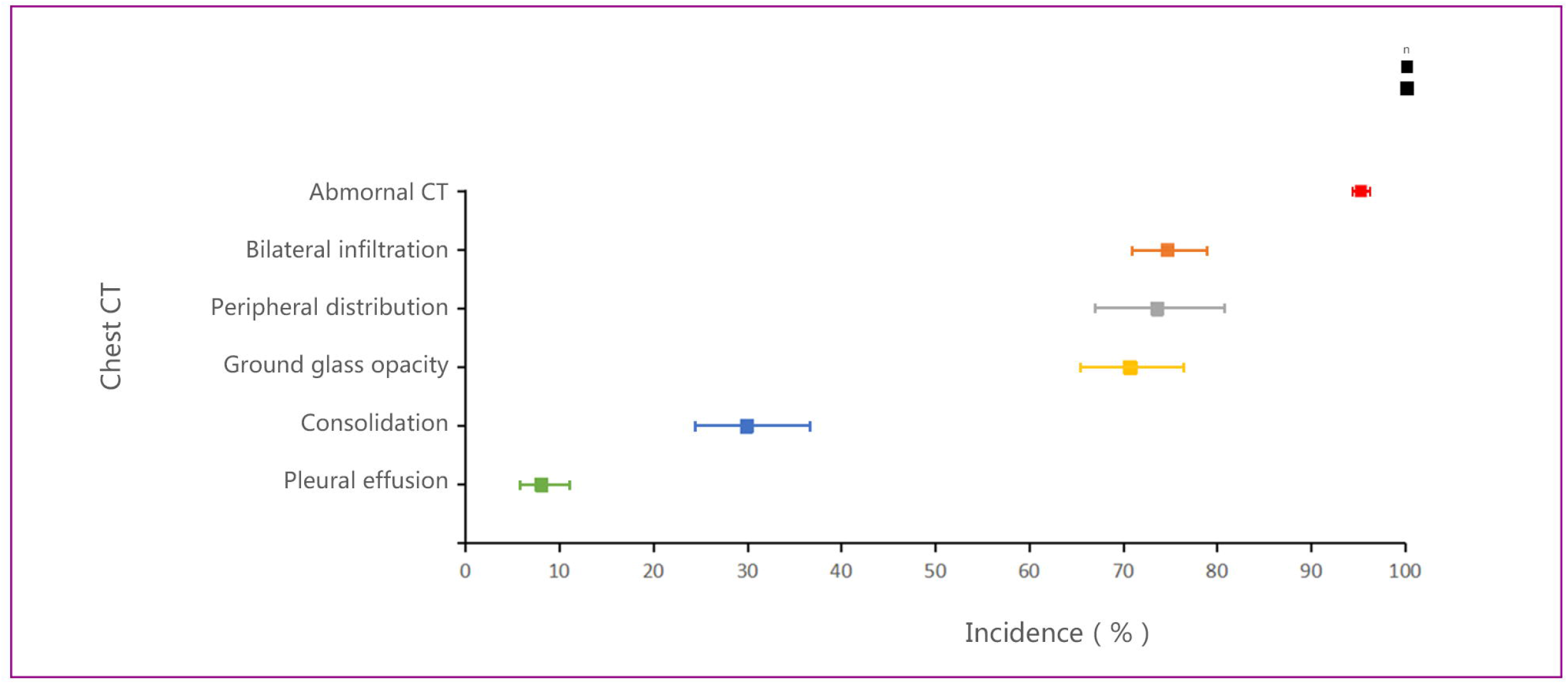
Estimates of the incidence of chest CT imaging for COVID-19 patients.

The mean values for the leucocyte count, neutrophil count, lymphocyte count, platelet count, and hemoglobin were all in the normal range. The proportion of patients with increased leukocytes and neutrophils were 10.55% and 4.05%, respectively, whereas those with decreased leukocytes, neutrophils, lymphocytes, platelets, and hemoglobin were 24.26%, 16.64%, 48.20%, 5.80%, and 23.56%, respectively.

The mean values for albumin, alanine transaminase (ALT), aspartate transaminase (AST), total bilirubin (TB), blood urea nitrogen (BUN), serum creatinine (SCr), creatine kinase (CK), and creatine kinase-MB (CK-MB) were all in the normal range. The mean value for lactate dehydrogenase (LDH) was moderately elevated to 308.76 U/L (95%CI 264.28–353.24 U/L). The proportion of patients showing decreased albumin was 37.51%, whereas the proportion of patients showing increased for ALT, AST, TB, BUN, SCr, LDH, CK, and CK-MB were 19.02%, 23.61%, 9.48%, 12.71%, 7.21%, 41.55%, 13.87%, and 16.76%, respectively. The mean values for potassium and sodium were in the normal range.

The mean values for activated partial thromboplastin time (APTT), prothrombin time (PT), D-Dimer, and fibrinogen were in the normal range. The proportion of patients showing increased APTT, PT, D-Dimer, and fibrinogen were 25.60%, 15.78%, 43.33%, and 36.90%, respectively.

Infection-related indicators including C-reactive protein (CRP), erythrocyte sedimentation rate (ESR), interleukin-6 (IL-6), and serum ferritin (SF) were all increased, with mean values of 32.05 mg/L (95%CI 27.34–36.75 mg/L), 36.37 mm/h (95%CI 30.53–42.21 mm/h), 13.90 pg/mL (11.53–16.26 pg/mL), and 714.73 ng/mL (95%CI 568.92–860.55ng/mL), respectively. Only procalcitonin (PCT) was in the normal range. The proportion of patients with increased CRP PCT, ESR, IL-6, and SF were 61.59%, 17.23%, 62.90%, 57.33%, and 76.47%, respectively. (Figure 5, Appendix pp 9-11)

**Figure 5.**
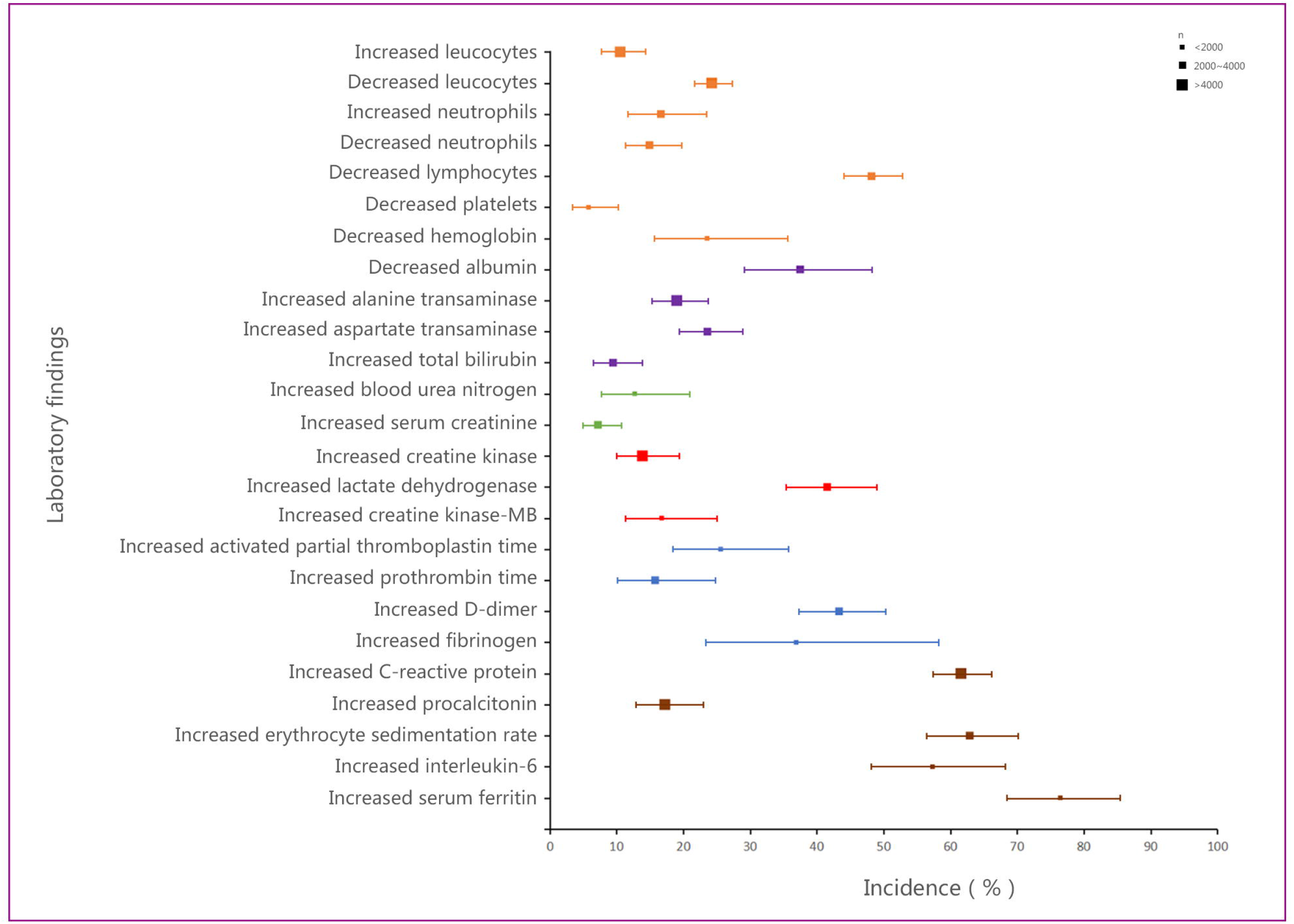
Estimates of the incidence of laboratory findings for COVID-19 patients.

### Complications and treatments of COVID-19

Common complications of COVID-19 included acute respiratory distress syndrome (ARDS) (25.09% 95%CI 18.88–3.35%), acute liver injury (21.68%, 95%CI 14.65–32.08%), secondary infection (17.14%, 95%CI 10.72–27.41%), acute cardiac injury (10.61%, 95%CI 7.20–15.64%), acute kidney injury (7.12%, 95%CI 4.74–10.70%), and shock (6.05%, 95%CI 3.09–11.85%). (see Figure 6, Appendix pp 12)

**Figure 6.**
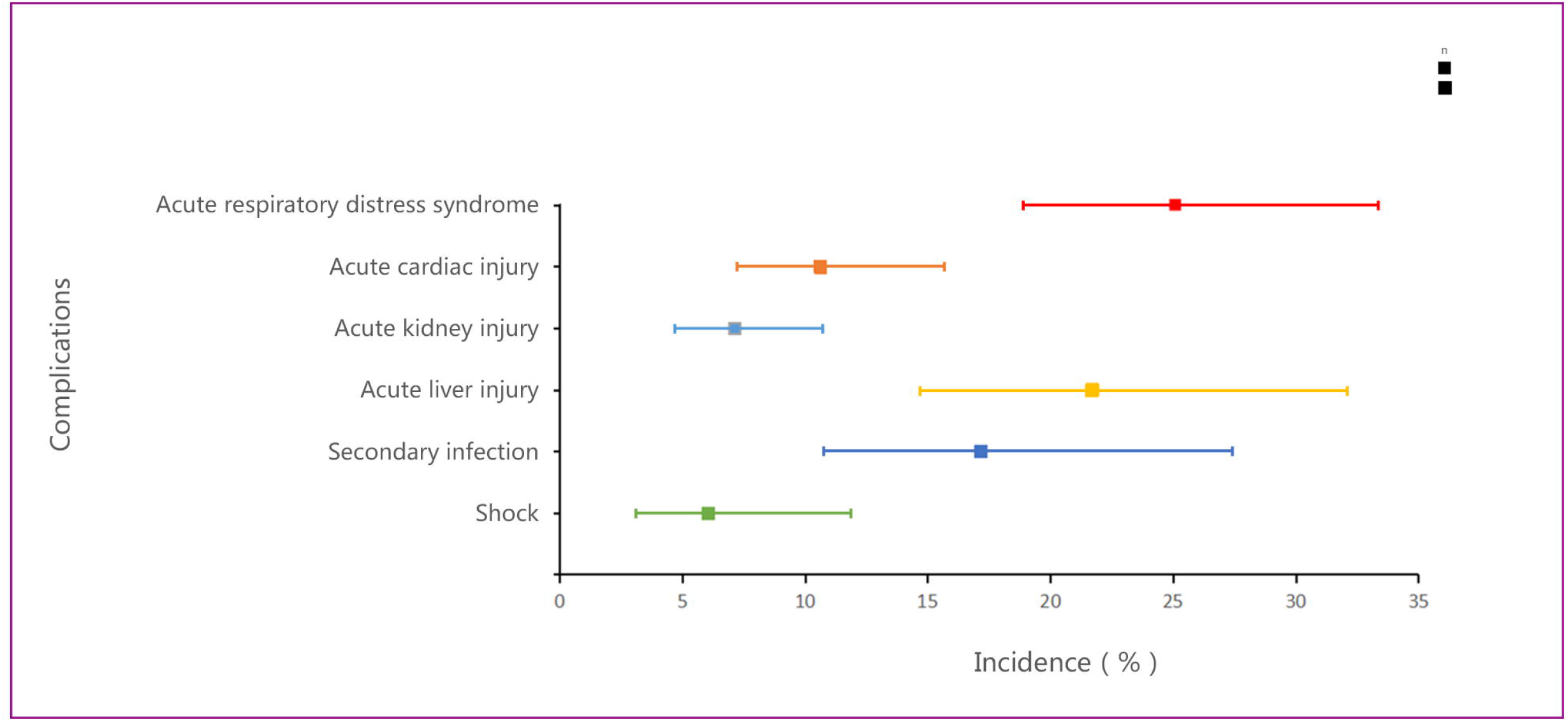
Estimates of the incidence of complications for COVID-19 patients.

Of 7510 COVID-19 patients, 5988 (89.75%, 95%CI 87.90–91.64%) were administered antiviral therapy, 79.13% with antibiotic therapy, 35.64% with corticosteroids, 31.69% with immunoglobulin, and 71.07% with γ-interferon. Additionally, 3073 of 5703 cases (61.91%) were administered oxygen support, 14.52% with non-invasive mechanical ventilation, and 7.48% with invasive mechanical ventilation. Sixty-two of 4012 cases (2.27%) were administered extracorporeal membrane oxygenation and 74 of 1656 cases (4.47%) were administered continuous renal replacement therapy. Further, 14.87% of patients with COVID-19 were admitted to the intensive care unit and 6262 patients reported clinical outcomes. There were 3664 hospitalizations; 2133 patients improved and were discharged, and 233 patients died. Mortality was 3.09% (1.85–5.15). (see Figure 7, Appendix 11-12)

**Figure 7.**
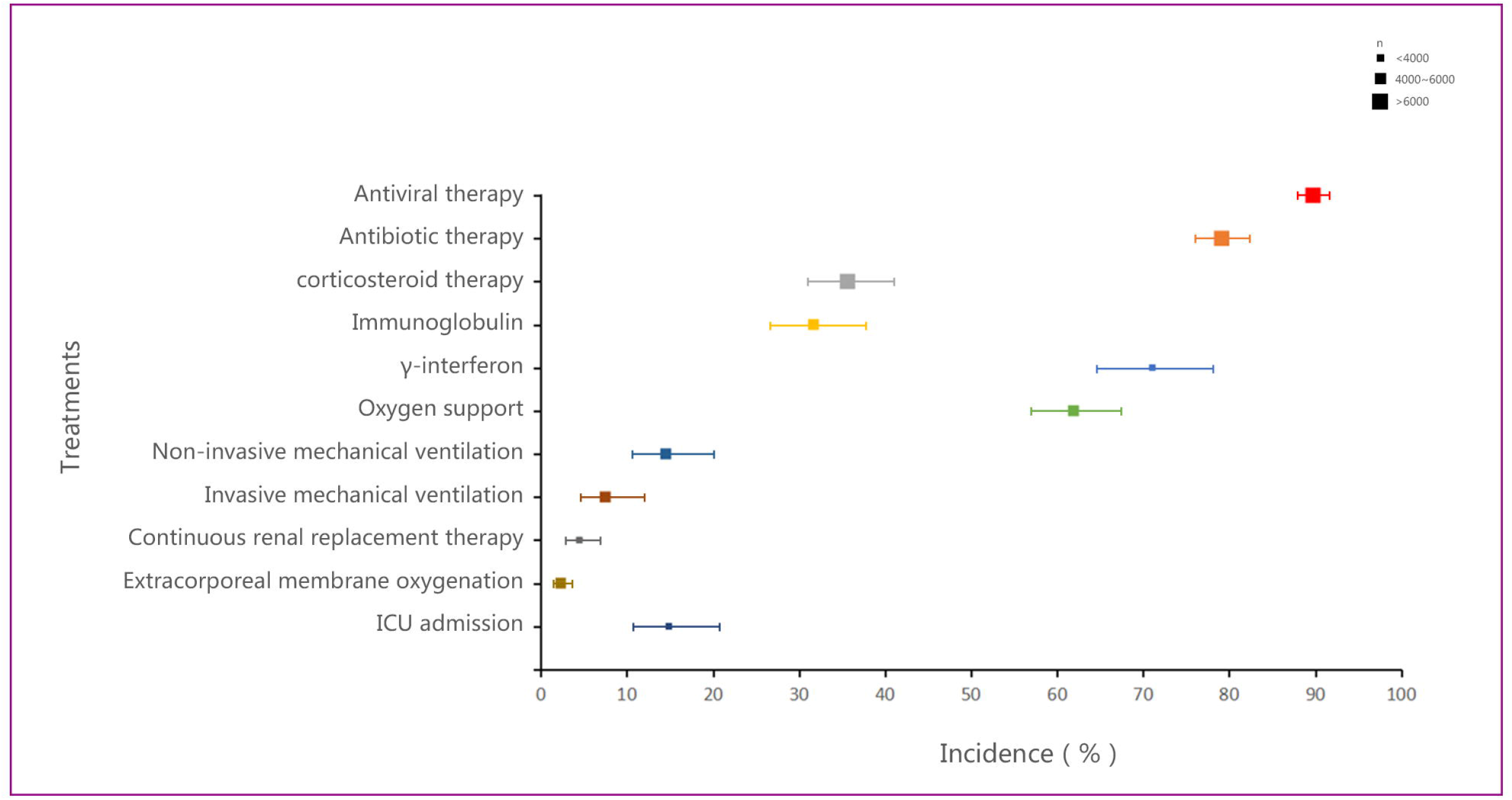
Estimates of the incidence of treatments for COVID-19 patients.

## Discussion

In this review, we retrospectively analyzed clinical data from patients with COVID-19. These patients were admitted to hospitals between December 2019 and February 2020, which covered the rapid growth period of the COVID-19 epidemic in mainland China. This meta-analysis is the first comprehensive summary of the clinical characteristics of COVID-19 in mainland China. Through literature retrieval and data extraction, we identified 20,662 patients confirmed to have COVID-19 reported in 147 articles. The patients ranged in age from 1 month to 100 years old, with a mean age was 49.40 years, which is consistent with a China CDC report.^157^ This suggests that the general population, regardless of age, is susceptible to SARS-CoV-2 infection. In addition, some studies suggested that advanced age is a risk factor for severe illness and mortality.^15,19,90,92^ The proportion of males was 53.45%, indicating that COVID-19 does not have a gender predisposition. Approximately 1 in 10 patients had a history of smoking, but whether this population was more susceptible to COVID-19 remains unclear. Of 8028 COVID-19 patients, 38.52% had at least one comorbidity, most commonly hypertension and diabetes. Any comorbidity is an important factor in poor prognosis. Patients with hypertension, diabetes, and cardiovascular and cerebrovascular diseases are at a higher risk of severe illness or death.^19,90^

We also summarized 20 clinical symptoms. The most common symptom observed in patients with COVID-19 was fever; approximately two-thirds of patients had mild to moderate fever, whereas only 11.45% of patients had high fever. The second most common symptom was cough, and approximately one-quarter of COVID-19 patients produced sputum. Because angiotensin-converting enzyme 2 of alveolar epithelial cells is the receptor for SARS-CoV-2,^158^ this infection induced excessive host immune responses, causing diffuse alveolar damage and lymphocytic infiltration in both lungs,^159^ manifested as dyspnea and shortness of breath. However, patients with COVID-19 rarely showed obvious signs and symptoms in the upper respiratory tract (pharyngalgia, rhinorrhea, nasal obstruction). Fatigue and myalgia were the most common musculoskeletal symptoms. In addition, gastrointestinal symptoms such as anorexia, diarrhea, nausea, vomiting, and abdominal pain were rare in patients with COVID-19. Thus, fever and cough are the most typical clinical symptoms.

In our study, most patients with COVID-19 had abnormal findings in chest CT imaging. The typical CT findings were characterized by ground glass opacity or consolidation. In addition, bilateral lungs infiltration and peripheral distribution were consistent with the pathological findings.^159^ According to the Guidelines for the Diagnosis and Treatment of Novel Coronavirus Infection, clinical diagnosis can be made by typical chest CT imaging, particularly when nucleic acid assays are scarce.^160^ In laboratory tests, the mean values of blood routine and biochemistry parameters were in the normal range. Nearly one-half of patients with COVID-19 had lymphocytopenia, and about a quarter of patients had leukopenia and anemia. Because angiotensin-converting enzyme 2 is widespread in the heart and liver, LDH was moderately elevated, and the proportion of patients showing increased for ALT, AST, CK, and CK-MB were 19.02%, 23.61%,13.87% and 16.76%, respectively. However, indicators of kidney function such as blood urea nitrogen, serum creatinine were not obviously elevated. In addition, the proportions of D-Dimer and fibrinogen were increased, suggesting that patients with COVID-19 had a fibrinolytic hyperactivity status. Infection-related indicators in patients with COVID-19 were generally increased, including the C-reactive protein, erythrocyte sedimentation rate, interleukin-6, and serum ferritin, suggesting the presence of an inflammatory storm or secondary infection in these patients. According to Wang et al.,^19^ patients with severe ill are more prone to laboratory abnormalities, including increased leukocytes, decreased lymphocytes, abnormal liver function, abnormal coagulation function, and increased infection-related indicators, leading to ARDS, acute myocardial injury, acute liver injury, and shock. Lymphocytopenia, increased LDH, increased D-dimer, and increased infection-related indicators, particularly IL-6, have been suggested to lead to a poor prognosis.^90,92^

Most patients with COVID-19 were administered antiviral therapy such as oseltamivir or lopinavir,^30,161^ whereas a large percentage of patients were administered γ-interferon as an anti-viral agent to improve immunity. Antibiotic therapy was used in most patients to treat secondary infection or sepsis. Additionally, anti-inflammatory treatment was essential because of the cytokine storm;^162^ more than one-third of patients were administered systemic corticosteroids. However, caution should be used when administering corticosteroids therapy avoid side-effects such as weakened immunity, nosocomial infections, psychosis, diabetes, and avascular necrosis.^163^ Only 233 deaths occurred among 6262 COVID-19 patients; the mortality rate was 3.09% consistent with data from the World Health Organization.^6^

This study had both strengths and limitations. This meta-analysis is the most comprehensive assessment and robust evidence to date of the clinical characteristics of patients with COVID-19 in mainland China. We evaluated 147 published and preprinted studies involving 20,662 patients, equivalent to a quarter of all confirmed cases in mainland China and reflecting the clinical characteristics of patients with COVID-19.^6^ There were some limitations to our study. First, this study was retrospective, and most studies were preprinted articles that had not been peer-reviewed, leading to a moderate overall quality of the literature. Second, not all clinical characteristics were well-documented, such as chest CT or laboratory tests, leading to inconsistencies in the total number of each item calculated. This also led to heterogeneity. Third, all included cases were treated from December 2019 to February 2020, and many were reported by several hospitals in Wuhan, Hubei province. There may be repeated reports of some cases, leading to a significant heterogeneity and obvious publication bias. To overcome these issues, we used random effects for meta-analysis, meta-regression tests, and Egger’s test. Finally, we were unable to determine the incubation period of COVID-19 because of the heterogeneity among studies in reporting the timeline of cases.

In summary, COVID-19 is an emerging infectious disease with various clinical manifestations. The morbidity of COVID-19 is not age- and sex-dependent. The most common clinical symptoms were fever (78.49%) and cough (55.49%). Chest CT imaging showed abnormal findings in most patients with ground glass opacity or consolidation around the periphery of the bilateral pulmonary. The main characteristics in laboratory findings were lymphopenia, increased LDH, increased infection-related indicators, and fibrinolytic hyperactivity. Complications such as ARDS and acute liver injury often occur in patients with severe ill. Antiviral therapy, antibiotics, and oxygen support are the most important treatments, and hormone therapy is used in some critical patients. Our findings provide important guidance for the current global pandemic in the diagnosis and treatment of COVID-19.

## Data Availability

All data referred to in the manuscript are available. All data in the submitted article.

## Contributors

JG, ZG, PY and CT conceived the idea of the study and developed the protocol. CT, KZ and ZL did the literature search and studies selection. CT, WW extracted relevant information. ZP and PY assessed the risk of bias in the eligible studies. CT synthesized the data and wrote the first draft of the manuscript. ZL, PY, ZG and JG revised successive drafts of the paper and approved the final version. ZG supervised the overall work and is the guarantor of the review.

## References

1 Zhu N, Zhang D, Wang W, et al. A novel coronavirus from patients with pneumonia in China, 2019. N Engl J Med 2020; 382:727–33.

2 Wu A, Peng Y, Huang B, et al. Genome Composition and Divergence of the Novel Coronavirus (2019-nCoV) Originating in China. Cell Host Microbe 2020; 27: 325–28.

3 Zhou P, Yang XL, Wang XG, et al. A pneumonia outbreak associated with a new coronavirus of probable bat origin. Nature 2020; 579: 270–73.

4 Lu R, Zhao X, Li J, et al. Genomic characterization and epidemiology of 2019 novel coronavirus: implications for virus origins and receptor binding. Lancet 2020; 395: 565–74.

5 World Health Organization. Naming the coronavirus disease (COVID-19) and the virus that causes it. https://www.who.int/emergencies/diseases/novel-coronavirus-2019/technical-guidance/naming-the-coronavirus-disease-(covid-2019)-and-the-virus-that-causes-it.

6 World Health Organization. Coronavirus disease (COVID-2019) situation reports-88. April 17, 2020. https://www.who.int/docs/default-source/coronaviruse/situation-reports/20200417-sitrep-88-covid-191b6cccd94f8b4f219377bff55719a6ed.pdf?sfvrsn=ebe78315_6 (accessed April 18, 2020).

7 Wan X, Wang W, Liu J, Tong T. Estimating the sample mean and standard deviation from the sample size, median, range and/or interquartile range. BMC Med Res Methodol 2014; 14: 135.

8 Luo D, Wan X, Liu J, Tong T. Optimally estimating the sample mean from the sample size, median, mid-range, and/or mid-quartile range. Stat Methods Med Res 2018; 27: 1785–805.

9 Sterne JA, Hernán MA, Reeves BC, et al. ROBINS-I: a tool for assessing risk of bias in non-randomised studies of interventions. BMJ 2016; 355: i4919.

10 Chen N, Zhou M, Dong X, et al. Epidemiological and clinical characteristics of 99 cases of 2019 novel coronavirus pneumonia in Wuhan, China: a descriptive study. Lancet 2020; 395: 507–13.

11 Liu K, Fang YY, Deng Y, et al. Clinical characteristics of novel coronavirus cases in tertiary hospitals in Hubei Province. Chin Med J (Engl) 2020; 00:00–00. DOI:10.1097/CM9.0000000000000744.

12 Huang C, Wang Y, Li X, et al. Clinical features of patients infected with 2019 novel coronavirus in Wuhan, China. Lancet 2020; 395: 497–506.

13 Wu J, Wu X, Zeng W, et al. Chest CT Findings in Patients with Corona Virus Disease 2019 and its Relationship with Clinical Features. Invest Radiol 2020; Published Online February 21. DOI:10.1097/RLI.0000000000000670.

14 Zhang JJ, Dong X, Cao YY, et al. Clinical characteristics of 140 patients infected with SARS-CoV-2 in Wuhan, China. Allergy 2020; 00:1–12. DOI:10.1111/all.14238.

15 Yang X, Yu Y, Xu J, et al. Clinical course and outcomes of critically ill patients with SARS-CoV-2 pneumonia in Wuhan, China: a single-centered, retrospective, observational study. Lancet Respir Med 2020; Published Online February 21. DOI:10.1016/S2213-2600(20)30079-5.

16 Xu XW, Wu XX, Jiang XG, et al. Clinical findings in a group of patients infected with the 2019 novel coronavirus (SARS-Cov-2) outside of Wuhan, China: retrospective case series. BMJ 2020; 368: m606. DOI:10.1136/bmj.m606.

17 Chung M, Bernheim A, Mei X, et al. CT Imaging Features of 2019 Novel Coronavirus (2019-nCoV). Radiology 2020; 295(1): 202–07.

18 Chang D, Lin M, Wei L, et al. Epidemiologic and Clinical Characteristics of Novel Coronavirus Infections Involving 13 Patients Outside Wuhan, China. N Engl J Med 2020; Published online February 7. DOI:10.1001/jama.2020.1623.

19 Wang D, Hu B, Hu C, et al. Clinical Characteristics of 138 Hospitalized Patients With 2019 Novel Coronavirus-Infected Pneumonia in Wuhan, China. JAMA 2020; Published online February 7. DOI:10.1001/jama.2020.1585.

20 Pan Y, Guan H, Zhou S, et al. Initial CT findings and temporal changes in patients with the novel coronavirus pneumonia (2019-nCoV): a study of 63 patients in Wuhan, China. Eur Radiol 2020; Published online February 13. DOI:10.1007/s00330-020-06731-x.

21 Shi H, Han X, Jiang N, et al. Radiological findings from 81 patients with COVID-19 pneumonia in Wuhan, China: a descriptive study. Lancet Infect Dis 2020; 20: 425–34.

22 Pan F, Ye T, Sun P, et al. Time Course of Lung Changes On Chest CT During Recovery From 2019 Novel Coronavirus (COVID-19) Pneumonia. Radiology 2020; 200370. DOI:10.1148/radiol.2020200370.

23 Liu M, He P, Liu H, et al. Clinical characteristics of 30 medical workers infected with new coronavirus pneumonia. Zhonghua Jie He He Hu Xi Za Zhi 2020;43(3):209–214.

24 Chen L, Liu H, Liu W, et al. Analysis of clinical features of 29 patients with 2019 novel coronavirus pneumonia. Zhonghua Jie He He Hu Xi Za Zhi 2020;43(3):203–208.

25 Feng K, Yun YX, Wang XF, et al. Analysis of CT features of 15 Children with 2019 novel coronavirus infection. Zhonghua Er Ke Za Zhi 2020;58(0):E007.

26 Liu C, Jiang ZC, Shao CX, et al. Preliminary study of the relationship between novel coronavirus pneumonia and liver function damage: a multicenter study. Zhonghua Gan Zang Bing Za Zhi 2020;28(2):148–152.

27 Cai J, Xu J, Lin D, et al. A Case Series of children with 2019 novel coronavirus infection: clinical and epidemiological features. Clin Infect Dis 2020; Published Online February 28. DOI:10.1093/cid/ciaa198.

28 Liu Y, Yang Y, Zhang C, et al. Clinical and biochemical indexes from 2019-nCoV infected patients linked to viral loads and lung injury. Sci China Life Sci 2020; 63: 364–74.

29 Yang W, Cao Q, Qin L, et al. Clinical characteristics and imaging manifestations of the 2019 novel coronavirus disease (COVID-19): A multi-center study in Wenzhou city, Zhejiang, China. J Infect 2020; Published Online February 26. DOI: 10.1016/j.jinf.2020.02.016.

30 Guan WJ, Ni ZY, Hu Y, et al. Clinical Characteristics of Coronavirus Disease 2019 in China. N Engl J Med 2020; Published Online March 2. DOI: 10.1056/NEJMoa2002032.

31 Wu J, Liu J, Zhao X, et al. Clinical Characteristics of Imported Cases of COVID-19 in Jiangsu Province: A Multicenter Descriptive Study. Clin Infect Dis 2020; Published Online February 29. DOI: 10.1093/cid/ciaa199.

32 Ai T, Yang Z, Hou H, et al. Correlation of Chest CT and RT-PCR Testing in Coronavirus Disease 2019 (COVID-19) in China: A Report of 1014 Cases. Radiology 2020; 200642. DOI: 10.1148/radiol.2020200642.

33 Song F, Shi N, Shan F, et al. Emerging 2019 Novel Coronavirus (2019-nCoV) Pneumonia. Radiology 2020; 295(1): 210–17.

34 Xu X, Yu C, Qu J, et al. Imaging and clinical features of patients with 2019 novel coronavirus SARS-CoV-2. Eur J Nucl Med Mol Imaging 2020; Published Online February 28. DOI: 10.1007/s00259-020-04735-9.

35 Lu HZ, Ai JW, Shen YZ, et al. A descriptive study of the impact of diseases control and prevention on the epidemics dynamics and clinical features of SARS-CoV-2 outbreak in Shanghai, lessons learned for metropolis epidemics prevention. medRxiv 2020; preprint. DOI: 10.1101/2020.02.19.20025031.

36 Fu HY, Li HJ, Tang XQ et al. Analysis on the Clinical Characteristics of 36 Cases of Novel Coronavirus Pneumonia in Kunming. medRxiv 2020; preprint. DOI: 10.1101/2020.02.28.20029173.

37 Yuan ML, Yin W, Tao ZW, et al. Association of radiologic findings with mortality of patients infected with 2019 novel coronavirus in Wuhan, China. medRxiv 2020; preprint. DOI: 10.1101/2020.02.22.20024927.

38 Jin JM, Bai P, He W, et al. Higher severity and mortality in male patients with COVID-19 independent of age and susceptibility. medRxiv 2020; preprint. DOI: 10.1101/2020.02.23.20026864.

39 Chen G, Wu D, Guo W, et al. Clinical and immunologic features in severe and moderate forms of Coronavirus Disease 2019. medRxiv 2020; preprint. DOI: 10.1101/2020.02.16.20023903.

40 Hu ZL, Song C, Xu CJ, et al. Clinical Characteristics of 24 Asymptomatic Infections with COVID-19 Screened among Close Contacts in Nanjing, China. medRxiv 2020; preprint. DOI: 10.1101/2020.02.20.20025619.

41 Li X, Wang LW, Yan SN, et al. Clinical characteristics of 25 death cases with COVID-19: a retrospective review of medical records in a single medical center, Wuhan, China. medRxiv 2020; preprint. DOI: 10.1101/2020.02.19.20025239.

42 Huang Y, Zhou HQ, Yang R, Xu Y, Feng XW, Gong P. Clinical characteristics of 36 non-survivors with COVID-19 in Wuhan, China. medRxiv 2020; preprint. DOI: 10.1101/2020.02.27.20029009.

43 Zhang BC, Zhou XY, Qiu YR, et al. Clinical characteristics of 82 death cases with COVID-19. medRxiv 2020; preprint. DOI: 10.1101/2020.02.26.20028191.

44 Liu L, Gao JY, Hu WM, et al. Clinical characteristics of 51 patients discharged from hospital with COVID-19 in Chongqing, China. medRxiv 2020; preprint. DOI: 10.1101/2020.02.20.20025536.

45 Cao WL, Shi L, Chen L, Xu XM, Wu ZR, et al. Clinical features and laboratory inspection of novel coronavirus pneumonia (COVID-19) in Xiangyang, Hubei. medRxiv 2020; preprint. DOI: 10.1101/2020.02.23.20026963.

46 Liu YL, Sun WW, Li J, et al. Clinical features and progression of acute respiratory distress syndrome in coronavirus disease 2019. medRxiv 2020; preprint. DOI: 10.1101/2020.02.17.20024166.

47 Cui PF, Chen Z, Wang T, et al. Clinical features and sexual transmission potential of SARS-CoV-2 infected female patients: a descriptive study in Wuhan, China. medRxiv 2020; preprint. DOI: 10.1101/2020.02.26.20028225.

48 Fan ZY, Chen LP, Li, J, et al. Clinical Features of COVID-19-Related Liver Damage. medRxiv 2020; preprint. DOI: 10.1101/2020.02.26.20026971.

49 Cai QX, Huang DL, Ou PC, et al. COVID-19 in a Designated Infectious Diseases Hospital Outside Hubei Province, China. medRxiv 2020; preprint. DOI: 10.1101/2020.02.17.20024018.

50 Qian GQ, Yang NB, Ding F, et al. Epidemiologic and Clinical Characteristics of 91 Hospitalized Patients with COVID-19 in Zhejiang, China: A retrospective, multi-centre case series. medRxiv 2020; preprint. DOI: 10.1101/2020.02.23.20026856.

51 Li J, Li SL, Cai YR, et al. Epidemiological and Clinical Characteristics of 17 Hospitalized Patients with 2019 Novel Coronavirus Infections Outside Wuhan, China. medRxiv 2020; preprint. DOI: 10.1101/2020.02.11.20022053.

52 Wu CM, Hu XL, Song JX, et al. Heart injury signs are associated with higher and earlier mortality in coronavirus disease 2019 (COVID-19). medRxiv 2020; preprint. DOI: 10.1101/2020.02.26.20028589.

53 Xu LD, Yuan J, Zhang YR, et al. Highland of COVID-19 outside Hubei: epidemic characteristics, control and projections of Wenzhou, China. medRxiv 2020; preprint. DOI: 10.1101/2020.02.25.20024398.

54 Chen D, Li XK, Song QF, et al. Hypokalemia and Clinical Implications in Patients with Coronavirus Disease 2019 (COVID-19). medRxiv 2020; preprint. DOI: 10.1101/2020.02.27.20028530.

55 Cheng YC, Luo R, Wang K, et al. Kidney impairment is associated with in-hospital death of COVID-19 patients. medRxiv 2020; preprint. DOI: 10.1101/2020.02.18.20023242.

56 Mao L, Wang MD, Chen SC, et al. Neurological Manifestations of Hospitalized Patients with COVID-19 in Wuhan, China: a retrospective case series study. medRxiv 2020; preprint. DOI: 10.1101/2020.02.22.20026500.

57 Yan L, Zhang HT, Xiao Y, et al. Prediction of survival for severe Covid-19 patients with three clinical features: development of a machine learning-based prognostic model with clinical data in Wuhan. medRxiv 2020; preprint. DOI: 10.1101/2020.02.27.20028027.

58 Liang Y, Liang JJ, Zhou QT, et al. Prevalence and clinical features of 2019 novel coronavirus disease (COVID-19) in the Fever Clinic of a teaching hospital in Beijing: a single-center, retrospective study. medRxiv 2020; preprint. DOI: 10.1101/2020.02.25.20027763.

59 Li J, Zhang YH, Wang F, et al. Sex differences in clinical findings among patients with coronavirus disease 2019 (COVID-19) and severe condition. medRxiv 2020; preprint. DOI: 10.1101/2020.02.27.20027524.

60 Ai JW, Chen JW, Wang Y, et al. The cross-sectional study of hospitalized coronavirus disease 2019 patients in Xiangyang, Hubei province. medRxiv 2020; preprint. DOI: 10.1101/2020.02.19.20025023.

61 Wang WJ, Liu XQ, Wu SP, et al. The definition and risks of Cytokine Release Syndrome-Like in 11 COVID-19-Infected Pneumonia critically ill patients: Disease Characteristics and Retrospective Analysis. medRxiv 2020; preprint. DOI: 10.1101/2020.02.26.20026989.

62 Lu JT, Hu SF, Fan R, et al. ACP Risk Grade: A Simple Mortality Index for Patients with Confirmed or Suspected Severe Acute Respiratory Syndrome Coronavirus 2 Disease (COVID-19) During the Early Stage of Outbreak in Wuhan, China. Available at SSRN: https://ssrn.com/abstract=3543603.

63 Liu WH, Wang F, Li G, et al. Analysis of 2019-nCoV Infection and Clinical Manifestations of Outpatients: An Epidemiological Study from the Fever Clinic in Wuhan, China. Available at SSRN: https://ssrn.com/abstract=3542153.

64 Peng MF, Yang J, Shi QX, et al. Artificial Intelligence Application in COVID-19 Diagnosis and Prediction. Available at SSRN: https://ssrn.com/abstract=3541119.

65 Tian S, Hu N, Lou J, et al. Characteristics of COVID-19 infection in Beijing. J Infect 2020; Published Online February 27. DOI: 10.1016/j.jinf.2020.02.018.

66 Ji MY, Yuan L, Shen W, et al. Characteristics of Disease Progress in 101 Hospitalized Patients with Corona Virus Disease 2019 in Wuhan, China. Available at SSRN: https://ssrn.com/abstract=3543620.

67 Wang F, Nie JY, Wang HL, et al. Characteristics of Peripheral Lymphocyte Subset Alteration in 2019-nCoV Pneumonia. Available at SSRN: https://ssrn.com/abstract=3539681.

68 Wang B, Liu, YB, Wang Y, et al. Characteristics of Pulmonary Auscultation in Patients with 2019 Novel Coronavirus in China. Available at SSRN: https://ssrn.com/abstract=3543593.

69 Qin XW, Qiu SH, Yuan YM, et al. Clinical Characteristics and Treatment of Patients Infected with COVID-19 in Shishou, China. Available at SSRN: https://ssrn.com/abstract=3541147.

70 Xie ZJ, Bao JF, Cai ZB, et al. Clinical Characteristics of 60 COVID-19-Infected Patients with or Without Renal Injury In Hangzhou, China. Available at SSRN: https://ssrn.com/abstract=3541126.

71 Ji D, Zhang DW, Chen Z, et al. Clinical Characteristics Predicting Progression of COVID-19. Available at SSRN: https://ssrn.com/abstract=3539674.

72 Zhang FQ, He L, Ouyang YL, et al. Clinical Features of 81 Hospitalized Patients with 2019 Novel Coronavirus-Infected Pneumonia in Jingzhou, China: A Descriptive Study. Available at SSRN: https://ssrn.com/abstract=3544834.

73 Lu CY, Yang WB, Hu YM, et al. Coronavirus Disease 2019 (COVID-19) Pneumonia: Early Stage Chest CT Imaging Features and Clinical Relevance. Available at SSRN: https://ssrn.com/abstract=3543606.

74 Liu XX, Jiang WL, Zeng Z, et al. Decreased Counts of T Lymphocyte Subsets Predict Prognosis in SARS-CoV-2-Infected Pneumonia in Wuhan, China: A Retrospective Study. Available at SSRN: https://ssrn.com/abstract=3544850.

75 Qin C, Zhou L, Hu Z, et al. Dysregulation of immune response in patients with COVID-19 in Wuhan, China. Clin Infect Dis 2020; Published Online Marah 12. DOI: 10.1093/cid/ciaa248.

76 Lin Y, Ji CL, Weng WD, et al. Epidemiological and Clinical Characteristics of 124 Elderly Outpatients with COVID-19 in Wuhan, China. Available at SSRN: https://ssrn.com/abstract=3543596.

77 Kuang YN, Zhang HQ, Zhou RZ, et al. Epidemiological and Clinical Characteristics of 944 Cases of 2019 Novel Coronavirus Infection of Non-COVID-19 Exporting City, Zhejiang, China. Available at SSRN: https://ssrn.com/abstract=3543604.

78 Zhang JJ, Yang SX, Xu Y, et al. Epidemiological and Clinical Characteristics of COVID-19 Infection Outside Wuhan, China: A Multicenter Study. Available at SSRN: https://ssrn.com/abstract=3546040.

79 Xiao KH, Huang M, Zhan FB, et al. Epidemiological and Clinical Features of 197 Patients Infected with 2019 Novel Coronavirus in Chongqing, China: A Single Center Descriptive Study. Available at SSRN: https://ssrn.com/abstract=3539687

80 Wu WX, Xu ZF, Jin YB, Pan AZ. Key Points of Clinical and CT Imaging Features of 2019 Novel Coronavirus (2019-nCoV) Imported Pneumonia Based on 21 Cases Analysis. Available at SSRN: https://ssrn.com/abstract=3543610.

81 Liu J, Li SM, Liu J, et al. Longitudinal Characteristics of Lymphocyte Responses and Cytokine Profiles in the Peripheral Blood of SARS-CoV-2 Infected Patients. Available at SSRN: https://ssrn.com/abstract=3539682.

82 Wang HJ, Luo SS, Shen Y, et al. Multiple Enzyme Release, Inflammation Storm and Hypercoagulability Are Prominent Indicators For Disease Progression In COVID-19: A Multi-Centered, Correlation Study with CT Imaging Score. Available at SSRN: https://ssrn.com/abstract=3544837.

83 Deng CH, Yang Y, Chen HW, et al. Ocular Dectection of SARS-CoV-2 in 114 Cases of COVID-19 Pneumonia in Wuhan, China: An Observational Study. Available at SSRN: https://ssrn.com/abstract=3543587.

84 Du RH, Liang LR, Yang CQ, et al. Patient Predisposition at Hospital Admission Indirectly Dictates Disease Severity, Clinical Course and Outcomes of COVID-19 Pneumonia Patients in Wuhan, China. Available at SSRN: https://ssrn.com/abstract=3543584.

85 Xie SS, Zhang GL, Yu H, et al. The epidemiologic and clinical features of suspected and confirmed cases of imported 2019 novel coronavirus pneumonia in North Shanghai, China. Available at SSRN: https://ssrn.com/abstract=3541125.

86 Liu L, Zhang DC, Tang SG, et al. The epidemiological and clinical characteristics of 2019 novel coronalvirus infection in Changsha, China. Available at SSRN: https://ssrn.com/abstract=3537093.

87 Su VY, Yang YH, Yang KY, et al. The Risk of Death in 2019 Novel Coronavirus Disease (COVID-19) in Hubei Province. Available at SSRN: https://ssrn.com/abstract=3539655.

88 Zhou B, She JQ, Wang YD, Ma XC. The Clinical Characteristics of Myocardial injury in Severe and Very Severe Patients with 2019 Novel Coronavirus Disease. Available at SSRN: https://ssrn.com/abstract=3539668.

89 Xia W, Shao J, Guo Y, Peng X, Li Z, Hu D. Clinical and CT features in pediatric patients with COVID-19 infection: Different points from adults. Pediatr Pulmonol 2020; Published Online March 5. DOI: 10.1002/ppul.24718.

90 Zhou F, Yu T, Du R, et al. Clinical course and risk factors for mortality of adult inpatients with COVID-19 in Wuhan, China: a retrospective cohort study. Lancet 2020; 395: 1054–62.

91 Li Y, Xia L. Coronavirus Disease 2019 (COVID-19): Role of Chest CT in Diagnosis and Management. AJR Am J Roentgenol 2020; 215:1–7.

92 Wu C, Chen X, Cai Y, et al. Risk Factors Associated With Acute Respiratory Distress Syndrome and Death in Patients With Coronavirus Disease 2019 Pneumonia in Wuhan, China. JAMA Intern Med 2020; Published Online March 13. DOI: 10.1001/jamainternmed.2020.0994.

93 Zhao W, Zhong Z, Xie X, Yu Q, Liu J. Relation Between Chest CT Findings and Clinical Conditions of Coronavirus Disease (COVID-19) Pneumonia: A Multicenter Study. AJR Am J Roentgenol 2020; 215:1–6.

94 Zhou S, Wang Y, Zhu T, Xia L. CT Features of Coronavirus Disease 2019 (COVID-19) Pneumonia in 62 Patients in Wuhan, China. AJR Am J Roentgenol 2020; 215:1–8.

95 Liu L, Liu WB, Zheng YQ, et al. A preliminary study on serological assay for severe acute respiratory syndrome coronavirus 2 (SARS-CoV-2) in 238 admitted hospital patients. medRxiv 2020; preprint. DOI: 10.1101/2020.03.06.20031856.

96 Tang AJ, Xu WH, Shen M, et al. A retrospective study of the clinical characteristics of COVID-19 infection in 26 children. medRxiv 2020; preprint. DOI: 10.1101/2020.03.08.20029710.

97 Xu HY, Hou KK, Xu H, et al. Acute Myocardial Injury of Patients with Coronavirus Disease 2019. medRxiv 2020; preprint. DOI: 10.1101/2020.03.05.20031591.

98 Liu R, Ming XY, Xu O, et al. Association of Cardiovascular Manifestations with In-hospital Outcomes in Patients with COVID-19: A Hospital Staff Data. medRxiv 2020; preprint. DOI: 10.1101/2020.02.29.20029348.

99 Chen ZW, Hu JJ, Zhang ZW, et al. Caution: The clinical characteristics of COVID-19 patients at admission are changing. medRxiv 2020; preprint. doi: https://doi.org/10.1101/2020.03.03.20030833.

100 Zhao ZH, Xie JJ, Yin M, et al. Clinical and Laboratory Profiles of 75 Hospitalized Patients with Novel Coronavirus Disease 2019 in Hefei, China. medRxiv 2020; preprint. DOI: 10.1101/2020.03.01.20029785.

101 Shi Q, Zhao KL, Yu J, et al. Clinical characteristics of 101 non-surviving hospitalized patients with COVID-19-A single center, retrospective study. medRxiv 2020; preprint. DOI: 10.1101/2020.03.04.20031039.

102 Wang YF, Zhou Y, Yang Z, Xia DP, Geng S. Clinical Characteristics of Patients with Severe Pneumonia Caused by the 2019 Novel Coronavirus in Wuhan, China. medRxiv 2020; preprint. DOI: 10.1101/2020.03.02.20029306.

103 Xu Y, Li YR, Zeng Q, et al. Clinical Characteristics of SARS-CoV-2 Pneumonia Compared to Controls in Chinese Han Population. medRxiv 2020; preprint. DOI: 10.1101/2020.03.08.20031658.

104 Zhang GQ, Hu C, Luo LJ, et al. Clinical features and outcomes of 221 patients with COVID-19 in Wuhan, China. medRxiv 2020; preprint. DOI: 10.1101/2020.03.02.20030452.

105 Cao M, Zhang DD, Wang YH, et al. Clinical Features of Patients Infected with the 2019 Novel Coronavirus (COVID-19) in Shanghai, China. medRxiv 2020; preprint. DOI: 10.1101/2020.03.04.20030395.

106 Xu YH, Xu ZH, Liu XS, et al. Clinical findings in critically ill patients infected with SARS-CoV-2 in Guangdong Province, China: a multi-center, retrospective, observational study. medRxiv 2020; preprint. DOI: 10.1101/2020.03.03.20030668.

107 Song CY, Xu J, He JQ, Lu YQ. COVID-19 early warning score: a multi-parameter screening tool to identify highly suspected patients. medRxiv 2020; preprint. DOI: 10.1101/2020.03.05.20031906.

108 Chen XH, Zhao BH, Qu YM, et al. Detectable serum SARS-CoV-2 viral load (RNAaemia) is closely associated with drastically elevated interleukin 6 (IL-6) level in critically ill COVID-19 patients. medRxiv 2020; preprint. DOI: 10.1101/2020.02.29.20029520.

109 Wang Y, Jiang WW, He Q, et al. Early, low-dose and short-term application of corticosteroid treatment in patients with severe COVID-19 pneumonia: single-center experience from Wuhan, China. medRxiv 2020; preprint. DOI: 10.1101/2020.03.06.20032342.

110 Liao JQ, Fan SB, Chen J, et al. Epidemiological and clinical characteristics of COVID-19 in adolescents and young adults. medRxiv 2020; preprint. DOI:10.1101/2020.03.10.20032136.

111 Chen X, Zheng F, Qing YH, et al. Epidemiological and clinical features of 291 cases with coronavirus disease 2019 in areas adjacent to Hubei, China: a double-center observational study. medRxiv 2020; preprint. DOI: 10.1101/2020.03.03.20030353.

112 Qi D, Yan XF, Tang XM, et al. Epidemiological and clinical features of 2019-nCoV acute respiratory disease cases in Chongqing municipality, China: a retrospective, descriptive, multiple-center study. medRxiv 2020; preprint. DOI: 10.1101/2020.03.01.20029397.

113 Yang PH, Ding YB, Xu Z, et al. Epidemiological and clinical features of COVID-19 patients with and without pneumonia in Beijing, China. medRxiv 2020; preprint. DOI: 10.1101/2020.02.28.20028068.

114 Yang Y, Shen CG, Li JX, et al. Exuberant elevation of IP-10, MCP-3 and IL-1 ra during SARS-CoV-2 infection is associated with disease severity and fatal outcome. medRxiv 2020; preprint. DOI: 10.1101/2020.03.02.20029975.

115 Zhu ZW, Tang JJ, Chai XP, et al. How to differentiate COVID-19 pneumonia from heart failure with computed tomography at initial medical contact during epidemic period. medRxiv 2020; preprint. DOI: 10.1101/2020.03.04.20031047.

116 Zeng Q, Li YZ, Huang G, et al. Mortality of COVID-19 is associated with cellular immune function compared to immune function in Chinese Han population. medRxiv 2020; preprint. DOI: 10.1101/2020.03.08.20031229.

117 Xing QS, Li GJ, Xing YH, et al. Precautions are needed for COVID-19 patients with coinfection of common respiratory pathogens. medRxiv 2020; preprint. DOI: 10.1101/2020.02.29.20027698.

118 Gao L, Jiang D, Wen XS, et al. Prognostic value of NT-proBNP in patients with severe COVID-19. medRxiv 2020; preprint. DOI: 10.1101/2020.03.07.20031575.

119 Chen XP, Ling JX, Mo PZ, et al. Restoration of leukomonocyte counts is associated with viral clearance in COVID-19 hospitalized patients. medRxiv 2020; preprint. DOI: 10.1101/2020.03.03.20030437.

120 Fan H, Zhang L, Huang B, et al. Retrospective analysis of clinical features in 101 death cases with COVID-19. medRxiv 2020; preprint. DOI: 10.1101/2020.03.09.20033068.

121 Li L, Li S, Xu MM, et al. Risk factors related to hepatic injury in patients with corona virus disease 2019. medRxiv 2020; preprint. DOI: 10.1101/2020.02.28.20028514.

122 Liu T, Zhang JY, Yang YH, et. The potential role of IL-6 in monitoring severe case of coronavirus disease 2019. medRxiv 2020; preprint. DOI: 10.1101/2020.03.01.20029769.

123 Qiu CF, Xiao Q, Liao X, et al. Transmission and clinical characteristics of coronavirus disease 2019 in 104 outside Wuhan patients, China. medRxiv 2020; preprint. DOI: 10.1101/2020.03.04.20026005.

124 Li JW, Long X, Luo HL, et al. Clinical characteristics of deceased patients infected with SARS-Cov-2 in Wuhan, China. Available at SSRN: https://ssrn.com/abstract=3546043.

125 Huang M, Yang Y, Shang FT, et al. Early and critical care in severe patients with COVID-19 in Jiangsu Province, China: a descriptive study. Available at SSRN: https://ssrn.com/abstract=3546056.

126 Shang J, Du RH, Lu QF, et al. The treatment and outcomes of patients with COVID-19 in Hubei, China: a multicentered, retrospective, observational study. Available at SSRN: https://ssrn.com/abstract=3546060.

127 Lu ZB, Chen M, Fan YZ, et al. Clinical Characteristics and Risk Factors for Fatal Outcome in Patients with 2019-Coronavirus Infected Disease (COVID-19) in Wuhan. Available at SSRN: https://ssrn.com/abstract=3546069.

128 Chen CM, Cao ML, Peng L, et al. Coronavirus Disease-19 Among Children outside Wuhan, China. Available at SSRN: https://ssrn.com/abstract=3546071.

129 Lin BL, Lei ZY, Cao HJ, et al. Comparison of epidemiological and clinical features of patients with coronavirus disease (COVID-19) in Wuhan and outside Wuhan, China. Available at SSRN: https://ssrn.com/abstract=3546082.

130 Du YZ, Tu L, Zhu PJ, et al. Clinical features of 85 fatal cases of COVID-19 from Wuhan: a retrospective observational study. Available at SSRN: https://ssrn.com/abstract=3546088.

131 Shi WY, Peng XQ, Liu TF, et al. Deep Learning-Based Quantitative Computed Tomography model in Predicting the Severity of COVID-19: A Retrospective Study in 196 Patients. Available at SSRN: https://ssrn.com/abstract=3546089.

132 Wang YH, Dong CJ, Li CG, et al. Temporal changes of CT findings in 90 patients with COVID-19 pneumonia: a longitudinal study. Available at SSRN: https://ssrn.com/abstract=3546091.

133 Du WJ, Yu JH, Wang H, et al. Clinical characteristics of COVID-19 in children compared with adults outside of Hubei Province in China. Available at SSRN: https://ssrn.com/abstract=3546097.

134 Wang K, Zuo PY, Liu YW, et al. Clinical and laboratory predictors of in-hospital mortality in 305 patients with COVID-19: a cohort study in Wuhan, China. Available at SSRN: https://ssrn.com/abstract=3546115.

135 Bai T, Tu SJ, Wei Y, et al. Clinical and laboratory factors predicting the prognosis of patients with COVID-19: an analysis of 127 patients in Wuhan, China. Available at SSRN: https://ssrn.com/abstract=3546118.

136 Wei XS, Wang X, Niu YR, et al. Clinical characteristics of SARS-CoV-2 infected pneumonia with diarrhea. Available at SSRN: https://ssrn.com/abstract=3546120.

137 Xu H, Huang SF, Liu SK, et al. Evaluation of the clinical characteristics of suspected or confirmed cases of COVID-19 during home care with isolation: A new retrospective analysis based on O2O. Available at SSRN: https://ssrn.com/abstract=3548746.

138 Liu SQ, Luo HY, Wang YC, Wang DL, Ju SH, Yang Y. Characteristics and associations with severity in COVID-19 patients: a multicenter cohort study from Jiangsu province, China. Available at SSRN: https://ssrn.com/abstract=3548753.

139 Liu DH, Li L, Wu X, et al. Pregnancy and perinatal outcomes of women with COVID-19 Pneumonia: a preliminary analysis. Available at SSRN: https://ssrn.com/abstract=3548758.

140 Li Y, Wang JS, Wang CT, et al. Characteristics of respiratory virus infection during the outbreak of 2019 novel coronavirus in Beijing. Available at SSRN: https://ssrn.com/abstract=3548768.

141 Wang GY, Wu CF, Zhang Q, et al. Epidemiological and Clinical Features of Corona Virus Disease 2019 (COVID-19) in Changsha, China. Available at SSRN: https://ssrn.com/abstract=3548770.

142 Chen JP, Wang X, Zhang ST, et al. Findings of acute pulmonary embolism in COVID-19 patients. Available at SSRN: https://ssrn.com/abstract=3548771.

143 Zhu X, Yuan WZ, Huang KS, et al. Clinical features and short-term outcomes of elderly patients with COVID-19 in Wuhan, China: a single-center, retrospective, observational study. Available at SSRN: https://ssrn.com/abstract=3548774.

144 Huang R, Zhu L, Xue LY, et al. Clinical findings of patients with coronavirus disease 2019 in Jiangsu province, China: a retrospective, multi-center study. Available at SSRN: https://ssrn.com/abstract=3548785.

145 Peng SK, Pan LG, Zhang SJ, et al. Imaging features in COVID-19 Patients: Analysis of Data from Patients in Non-pandemic areas. Available at SSRN: https://ssrn.com/abstract=3548786.

146 Ma HJ, Hu JN, Tian J, et al. Visualizing the novel coronavirus (COVID-19) in children: What we learn from patients at Wuhan Children’s Hospital. Available at SSRN: https://ssrn.com/abstract=3550012.

147 Li X, Hu Y, Zhu SY, et al. Epidemiological feature and outcome of 292 hospitalized patients with COVID-19 under adequate medical resource condition. Available at SSRN: https://ssrn.com/abstract=3550016.

148 Duan QH, Guo GY, Ren Y, et al. Treatment Outcomes, Influence Factors of 116 Hospitalized COVID-19 Patients with Longer/Prolonged Treatment Course in Wuhan, China. Available at SSRN: https://ssrn.com/abstract=3550017.

149 Yao T, Gao Y, Cui Q, et al. Clinical characteristics of 55 cases of deaths with COVID-19 pneumonia in Wuhan, China: retrospective case series. Available at SSRN: https://ssrn.com/abstract=3550019.

150 Li YN, Wang MD, Zhou YF, et al. Acute cerebrovascular disease following COVID-19: a single center, retrospective, observational study. Available at SSRN: https://ssrn.com/abstract=3550025.

151 Mao B, Liu Y, Chai YH, et al. Early Discern COVID-19 from the Suspected Patients via Fever Clinics: A Multicenter Cohort Study from Shanghai. Available at SSRN: https://ssrn.com/abstract=3550037.

152 Wang Y, Yao L, Zhang JP, et al. Clinical characteristics and laboratory indicator analysis of 69 COVID-19 pneumonia patients in Suzhou, China. Available at SSRN: https://ssrn.com/abstract=3550041.

153 Hu XF, Zeng WB, Zhang YH, et al. CT Imaging Features of Different Clinical Types of COVID-19: A Chinese Multicenter Study. Available at SSRN: https://ssrn.com/abstract=3550043.

154 Wu Y, Hou JB, Wan X, et al. Epidemiologic and clinical characteristics of surgical patients infected with COVID-19 in Wuhan. Available at SSRN: https://ssrn.com/abstract=3550044.

155 Zhang Q, Shi HS, Meng DQ, et al. CT image guided handling of pneumonia patients with negative RT-PCR detection of SARS-CoV-2 in high epidemic areas. Available at SSRN: https://ssrn.com/abstract=3550061.

156 Yu HT, Li DY, Deng ZG, et al. Total protein as a biomarker for predicting coronavirus disease-2019 pneumonia. Available at SSRN: https://ssrn.com/abstract=3551289.

157 Wu Z, McGoogan JM. Characteristics of and Important Lessons From the Coronavirus Disease 2019 (COVID-19) Outbreak in China: Summary of a Report of 72LJ314 Cases From the Chinese Center for Disease Control and Prevention. JAMA 2020. Published online February 24. doi: 10.1001/jama.2020.2648.

158 Hoffmann M, Kleine-Weber H, Schroeder S, et al. SARS-CoV-2 Cell Entry Depends on ACE2 and TMPRSS2 and Is Blocked by a Clinically Proven Protease Inhibitor. Cell 2020. DOI: 10.1016/j.cell.2020.02.052.

159 Xu Z, Shi L, Wang Y, et al. Pathological findings of COVID-19 associated with acute respiratory distress syndrome. Lancet Respir Med 2020; published online February 17. DOI: 10.1016/S2213-2600(20)30076-X.

160 National Health Commission of the People’s Republic of China. Guidelines for the Diagnosis and Treatment of COVID-19 (Trial Version 7). March 4, 2020. http://www.nhc.gov.cn/yzygj/s7653p/202003/46c9294a7dfe4cef80dc7f5912eb1989.shtml (accessed March 5, 2020)

161 Cao B, Wang Y, Wen D, et al. A Trial of Lopinavir-Ritonavir in Adults Hospitalized with Severe Covid-19. N Engl J Med 2020. published on March 18. DOI: 10.1056/NEJMoa2001282.

162 Stebbing J, Phelan A, Griffin I, et al. COVID-19: combining antiviral and anti-inflammatory treatments. Lancet Infect Dis 2020; 20: 400–02.

163 Shang L, Zhao J, Hu Y, Du R, Cao B. On the use of corticosteroids for 2019-nCoV pneumonia. Lancet 2020; 395: 683–84.

